# Detection of mpox clade Ib nucleic-acids in wastewater solids at 147 wastewater treatment plants across the United States

**DOI:** 10.1101/2025.02.19.25322452

**Authors:** Alexandria B. Boehm, Marlene K. Wolfe, Amanda L. Bidwell, Bradley J. White, Bridgette Shelden, Dorothea Duong

**Affiliations:** Department of Civil and Environmental Engineering, Stanford University, 473 Via Ortega, Stanford, California, 94305; Gangarosa Department of Environmental Health, Rollins School of Public Health, Emory University, 1518 Clifton Rd, Atlanta, GA, USA, 30322; Verily Life Sciences LLC, South San Francisco, CA, USA; Google LLC, Mountain View, CA, USA

## Abstract

We used a sensitive, specific PCR assay to detect mpox clade Ib DNA in over 3000 wastewater samples collected prospectively across the United States. Mpox clade Ib DNA was detected in one sample from a location with a confirmed case; it was not detected in locations with no confirmed cases.

## Introduction

There are two clades of the mpox virus: clade I (subclades Ia and Ib) and clade II^1^. Mpox clade II is endemic to West Africa but began to spread across the world in 2022, causing a global outbreak^1^. Mpox clade I (now Ia) is endemic in Central Africa with outbreaks primarily in children,^1^ but in 2024, a new sub-clade, clade Ib, was identified in the Democratic Republic of Congo^1^. Clade Ib primarily infects adults and has spread rapidly since its emergence primarily through sexual contact^1^. As of mid-February 2025, there have been 100,000 cases of mpox Ib in 122 countries around the globe^2^. As of 12 February 2025, there have been four confirmed travel-associated mpox clade Ib cases in the United States (US); one in California on 15 November 2024^3^, Georgia on 14 January 2025^2^, New Hampshire on 7 February 2025^4^, and one in New York on 11 February 2025^5^. Outbreaks of clade I have previously been associated with a higher case fatality rate than clade II, but data to establish the severity of clade Ib during the current outbreak is limited^1^.

In previous work, we established that mpox clade II DNA can be detected in wastewater solids, and its concentrations correlated to community incident cases of infections^6^. We further showed that mpox clade II DNA preferentially partitions to solids in wastewater^6^. Mpox clade II DNA is shed in saliva, feces, urine, semen, and/or skin lesions of infected individuals^7^; DNA from these may end up in wastewater^8^. We have been prospectively measuring mpox clade II DNA across the US for use by public health to identify outbreaks^9^.

## The Study

In this study, we aimed to test whether mpox clade Ib DNA can be detected in wastewater solids to provide information on potential spread. Recent work has established that mpox clade I DNA is shed via excretions^10^ that enter wastewater.

We used a previously published and validated mpox clade 1b hydrolysis-probe PCR assay^11^ (Table 1) in a droplet digital PCR format. We confirmed the specificity and sensitivity of the assay in vitro and in silico (see appendix).

**Table 1.**
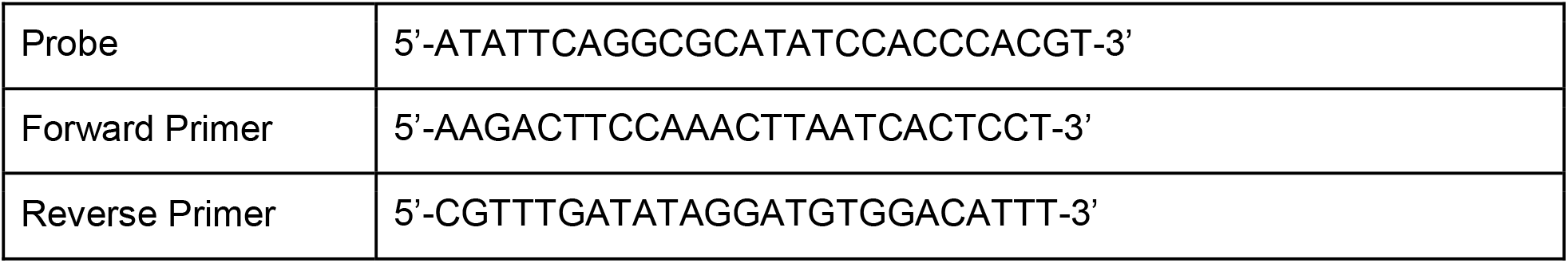
Mpox Clade Ib primers and probes^15^.

We retrospectively applied the assay to ten wastewater samples using methods described in the appendix. The ten samples were collected from a WWTP serving 750,000 people in San Francisco, California, USA. Mpox clade II DNA was previously detected at both high and low concentrations in these samples and the samples were collected at a time when mpox clade Ib was not circulating in the United States (see appendix for exact day and previously measured and reported mpox clade II concentrations). The samples were stored at -80°C as extracted and purified nucleic-acids for between one and three years prior to analysis. The nucleic-acids were obtained using the same methods as described below for the samples collected prospectively.

We confirmed limited degradation of nucleic-acids during storage by also measuring concentrations of the N gene from SARS-CoV-2, and comparing the measurements to those obtained from the same samples before storage (see appendix). Mpox clade Ib DNA concentrations were non-detectable in the ten samples, supporting the specificity of the molecular assay.

We then began prospective measurements of mpox clade 1b DNA in samples from 147 WWTPs located in 40 states (Figure 1). Samples were collected between 22 November 2024 and 31 January 2025, typically three times per week. A full description of WWTP locations and the number of samples and the time during which they were collected are provided in the appendix. Between 11 and 71 samples were tested per WWTP (median = 21 per WWTP).

**Figure 1.**
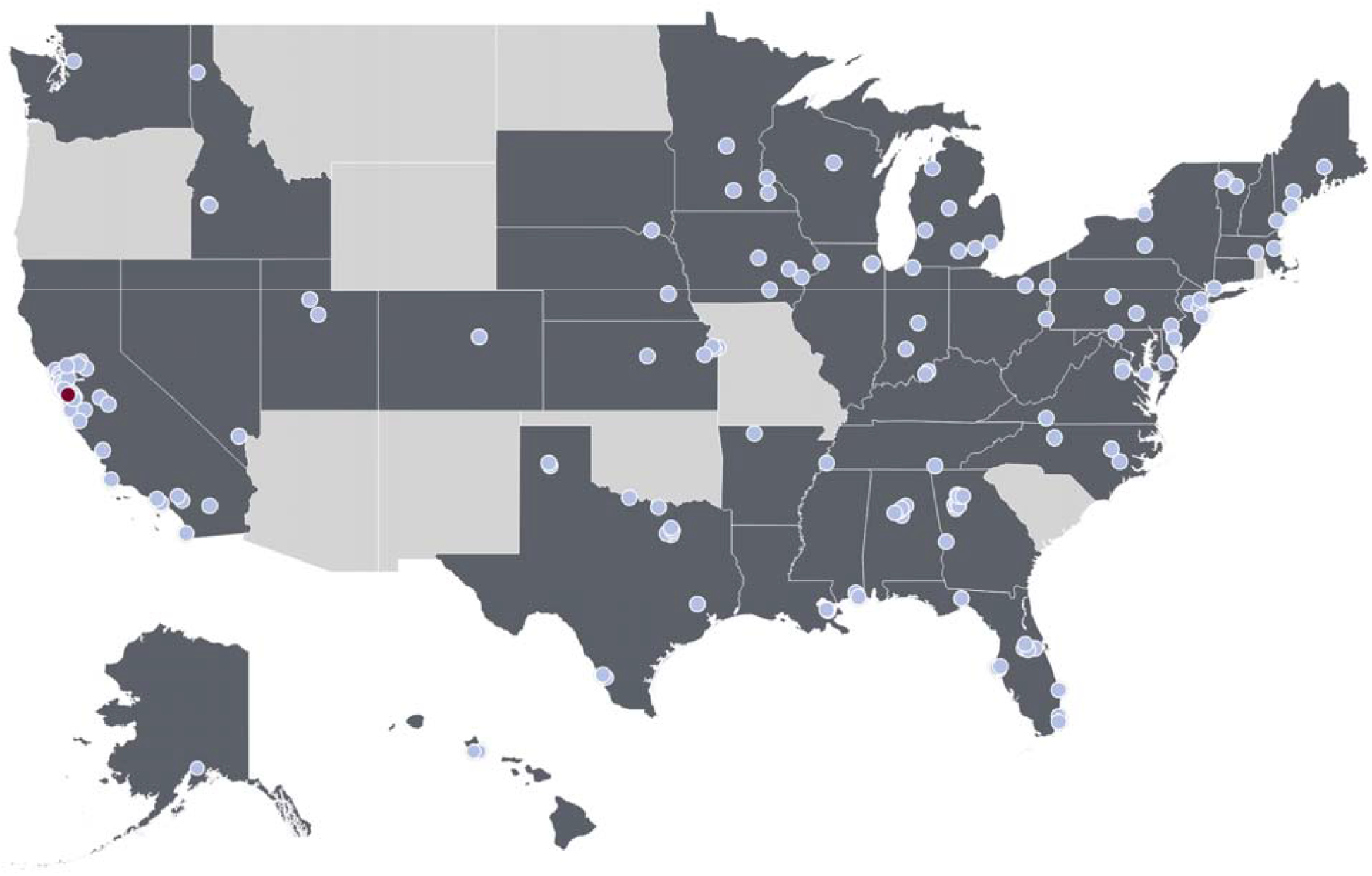
Map of the wastewater treatment plants (WWTPs) enrolled in this study. States where there are enrolled sites are shaded in dark gray. The red dot represents the location of the WWTP where there was a positive detection of mpox clade Ib (n=1), and the light blue dots represent participating WWTP where all samples were non-detect (n=146).

Sampling methods are provided in the appendix and in other publications^9^. After collection, samples were stored at 4 ◻ °C and shipped to the laboratory on ice and processed to completion within 48 hours of receipt at the laboratory. We do not expect the time between sample collection and analysis to affect target quantification as our work and others ^12–14^ show limited decay of the short length nucleic-acid targets over weeks at 4°C. A total of 3292 samples were collected and analyzed for this study.

Samples were processed and their nucleic acids extracted and purified from wastewater solids using the methods in the appendix and previously published^9^. Concentrations of mpox clade Ib DNA were measured using droplet digital PCR on 6-10 replicates for each sample. Extraction and PCR positive and negative controls were run on each 96-well plate. Results from replicate wells were merged for analysis. Concentrations are presented as copies per gram dry weight. For a sample to be scored as a positive, there had to be at least 3 positive droplets. The lowest measurable concentration is approximately 1000 copies/g dry weight (corresponds to three positive droplets). Errors are reported as standard deviations of the measurements as obtained from QX Manager Software (Bio-Rad, version 2.0). Data are available through the Stanford Digital Repository (https://doi.org/10.25740/vk458br0887).

Only one sample of 3292 tested positive. In the positive sample, the concentration of mpox clade 1b DNA was 1174 copies per gram dry weight of solids (cp/g, 68% confidence range between 674 and 1875 cp/g). The positive sample was obtained from a sewershed in California serving 199,000 individuals including an individual who had a confirmed infection of mpox clade Ib^3^. The positive sample was collected on 16 December 2024, 29 days after a news story broke of a travel-associated infection in the community. Mpox clade I DNA shedding from infected patients via skin, feces, saliva, and mucus can persist for over 20 days post symptom onset^10^. It is therefore possible that DNA from excretions of the infected individual lead to the positive detection.

Three thousand two hundred and ninety one (3291) samples were negative for mpox clade 1b DNA, consistent with a lack of known cases across the US. A negative sample is defined as a sample with less than three positive droplets across all generated droplets for that sample.

Negative controls and positive controls in this study performed as expected where they were negative and positive, respectively. Additional details are provided in the appendix. There have also been confirmed cases of mpox clade Ib infections in Georgia, New Hampshire, and New York. However, according to popular news stories these individuals were not expected to be within a WWTP sewershed included in this study, and the New Hampshire and New York cases were first reported after the last sample was collected.

## Conclusions

This study establishes methods for measuring mpox clade Ib DNA in wastewater solids for surveillance of mpox clade Ib infections. Results from analysis of wastewater solids samples are consistent with information on the presence and absence of mpox clade Ib infections in the included WWTP sewersheds during the study, and suggest wastewater monitoring for mpox clade Ib may be useful to monitor disease outbreaks.

Only one sample was positive for mpox clade Ib DNA and that sample was from a location where there was a known, travel-associated case. Samples from the same location prior to, and after, the positive samples were negative and there were no secondary cases reported.

Variability in concentrations of rare nucleic-acid targets in wastewater should be expected, and is caused by factors separate from those causing variability in clinical case or syndromic data. Mpox clade Ib DNA concentrations in wastewater solids may vary temporally owing to temporal variability in shedding by infected individuals, movement of people into and out of the sewershed, or deliveries of septic wastes to the system. Additionally wastewater solids may have variable concentrations spatially within the sampled material - spatial heterogeneities in the solids may contain variable amounts of mpox clade Ib DNA. To address variability, we processed high frequency samples and ran 6-10 replicates and numerous controls to ensure high quality measurements.

## Data Availability

Data are available through the Stanford Digital Repository (https://doi.org/10.25740/vk458br0887).

https://doi.org/10.25740/vk458br0887

## Acknowledgment

We thank the participating wastewater treatment plants for their samples for the project.

## References

(1) Titanji, B. K.; Hazra, A.; Zucker, J. Mpox Clinical Presentation, Diagnostic Approaches, and Treatment Strategies: A Review. JAMA 2024, 332 (19), 1652–1662. 10.1001/jama.2024.21091.

(2) United States Center for Disease Control. Mpox in the United States and Around the World: Current Situation. https://www.cdc.gov/mpox/situation-summary/index.html (accessed 2025-02-05).

(3) United States Center for Disease Control and Prevention Health Alert Network. First Case of Clade I Mpox Diagnosed in the United States. https://www.cdc.gov/han/2024/han00519.html (accessed 2025-02-05).

(4) New Hampshire Department of Health and Human Services. DHHS Identifies New Hampshire Resident with Clade I Mpox. https://www.dhhs.nh.gov/news-and-media/dhhs-identifies-new-hampshire-resident-clade-i-mpox (accessed 2025-02-11).

(5) New York State Department of Health. New York State Department of Health Issues Health Advisory on First Mpox Clade Ib Case Detected in New York. https://www.health.ny.gov/press/releases/2025/2025-02-12_mpox.htm (accessed 2025-02-18).

(6) Wolfe, M. K.; Yu, A. T.; Duong, D.; Rane, M. S.; Hughes, B.; Chan-Herur, V.; Donnelly, M.; Chai, S.; White, B. J.; Vugia, D. J.; Boehm, A. B. Use of Wastewater for Mpox Outbreak Surveillance in California. N Engl J Med 2023. 10.1056/NEJMc2213882.

(7) Peiró-Mestres, A.; Fuertes, I.; Camprubí-Ferrer, D.; Marcos, M. Á.; Vilella, A.; Navarro, M.; Rodriguez-Elena, L.; Riera, J.; Català, A.; Martínez, M. J.; Blanco, J. L. Frequent Detection of Monkeypox Virus DNA in Saliva, Semen, and Other Clinical Samples from 12 Patients, Barcelona, Spain, May to June 2022. Euro Surveill 2022, 27 (28). 10.2807/1560-7917.ES.2022.27.28.2200503.

(8) Chen, W.; Bibby, K. Model-Based Theoretical Evaluation of the Feasibility of Using Wastewater-Based Epidemiology to Monitor Monkeypox. Environ. Sci. Technol. Lett. 2022, 9 (9), 772–778. 10.1021/acs.estlett.2c00496.

(9) Boehm, A. B.; Wolfe, M. K.; Bidwell, A. L.; Zulli, A.; Chan-Herur, V.; White, B. J.; Shelden, B.; Duong, D. Human Pathogen Nucleic Acids in Wastewater Solids from 191 Wastewater Treatment Plants in the United States. Scientific Data 2024, 11 (1), 1141. 10.1038/s41597-024-03969-8.

(10) Yang, Y.; Niu, S.; Shen, C.; Yang, L.; Song, S.; Peng, Y.; Xu, Y.; Guo, L.; Shen, L.; Liao, Z.; Liu, J.; Zhang, S.; Cui, Y.; Chen, J.; Chen, S.; Huang, T.; Wang, F.; Lu, H.; Liu, Y. Longitudinal Viral Shedding and Antibody Response Characteristics of Men with Acute Infection of Monkeypox Virus: A Prospective Cohort Study. Nature Communications 2024, 15 (1), 4488. 10.1038/s41467-024-48754-8.

(11) Schuele, L.; Masirika, L. M.; Udahemuka, J. C.; Siangoli, F. B.; Mbiribindi, J. B.; Ndishimye, P.; Aarestrup, F. M.; Koopmans, M.; Oude Munnink, B. B.; Molenkamp, R.; group, G. M. Real-Time PCR Assay to Detect the Novel Clade Ib Monkeypox Virus, September 2023 to May 2024. Eurosurveillance, 2024, 29, 2400486.

(12) Roldan-Hernandez, L.; Graham, K. E.; Duong, D.; Boehm, A. B. Persistence of Endogenous SARS-CoV-2 and Pepper Mild Mottle Virus RNA in Wastewater-Settled Solids. ACS EST Water 2022. 10.1021/acsestwater.2c00003.

(13) Zhang, M.; Roldan-Hernandez, L.; Boehm, A. B. Persistence of Human Respiratory Viral RNA in Wastewater-Settled Solids. Applied and Environmental Microbiology 2024, 90 (4), e02272–23. 10.1128/aem.02272-23.

(14) Muirhead, A.; Zhu, K.; Brown, J.; Basu, M.; Brinton, M. A.; Costa, F.; Hayat, M. J.; Stauber, C. E. Zika Virus RNA Persistence in Sewage. Environ. Sci. Technol. Lett. 2020. 10.1021/acs.estlett.0c00535.

(15) Schuele, L.; Masirika, L. M.; Udahemuka, J. C.; Siangoli, F. B.; Mbiribindi, J. B.; Ndishimye, P.; Aarestrup, F. M.; Koopmans, M.; Munnink, B. B. O.; Molenkamp, R.; Group, G. M. Real-Time PCR Assay to Detect the Novel Clade Ib Monkeypox Virus, September 2023 to May 2024. Eurosurveillance 2024, 29 (32), 2400486. 10.2807/1560-7917.ES.2024.29.32.2400486.

