## Appendix for "Detection of mpox clade Ib nucleic-acids in wastewater solids at 147 wastewater treatment plants across the United States"

### Methods

**Assay choice.** A previously published assay[1] was used to detect clade Ib mpox virus genomic DNA in wastewater (Table 1, main text). This assay was designed in response to the emergence of clade Ib, including the introduction of a deletion in the complement binding protein (C3L) gene which has been commonly used to identify clade I mpox viruses. The assay spans the deletion; targeting regions upstream and downstream of the C3L deletion in Clade Ib. The original authors validated the assay by testing clinical samples from the clade Ib outbreak in the Democratic Republic of Congo[1].

**In silico assay testing.** We downloaded mpox genome sequences from GISAID on 20 August 2024 to confirm that the region targeted by the assay matches recent clade Ib sequences available at the time (2 from Kenya, 2 from Uganda, and 32 from DRC for Clade Ib). The primers and probe sensitivity and specificity were also tested using National Center for Biotechnology Information (NCBI) BLAST. No cross reactivity was identified.

**In vitro assay testing.** The assay was tested in vitro for sensitivity and specificity against extracted nucleic acids from respiratory viruses and non-target nucleic acids (Table S1). Nucleic-acids were extracted and purified as described below for the wastewater solids samples and then used neat as template in droplet digital 1-step RT-PCR assays. Assays were run in a single well using the cycling conditions and post processing using a droplet reader as for the wastewater samples in singleplex. No non-target samples produced positive droplets.

To test the quantitative sensitivity of the assay, we challenged the assay to serial dilutions of a positive control. The positive control was a synthetic nucleic acid gene block purchased from IDT (Coralville, Iowa) with the sequence:
ATTAGAATTTTCTATTTCAACGGGTATAGCAGAATATTTGAAACACGGCACTTCGAAATGGAAA
AGACTTCCAAACTTAATCACTCCTAGATATTCAGGCGCATATCCACCCACGTGTCAGATTGTT
AAATGTCCACATCCTATATCAAACGGAAAACTTCTAGCGGCTTAAAAGATCATACACTCATACA
ACGACAATGTAGACTTTAAGTGC.

The positive control was serially diluted and then 1.1 µL of each dilution was spiked into 4.4 µL of wastewater solids nucleic-acid extracts known to be negative for the target (collected prior to outbreak started using methods described below) to produce a total of 5.5 µL of template. The template was used in the PCR reactions as described below. We ran the PCR with and without the RT step to confirm that the inclusion of the RT step would not bias the results. As our project detects mostly RNA viruses, it is economical to use RT-PCR for all targets if possible. Inclusion of the RT step did not affect the quantification of the target (paired t-test, p=0.09) (Figure S2). The assay detected down to at least 2 copies per µl reaction.

**Wastewater sample collection for retrospective study.** Wastewater samples were obtained from a wastewater treatment plant (WWTP) located in the San Francisco Bay Area serving approximately 750,000 people (SEP). Samples were collected using sterile technique in clean, labeled bottles, and were "grab" samples from the primary clarifier. More details about the site are available elsewhere[2]. We selected samples from 7 days in August 2022 and 3 days in September - October 2024, the majority of which had quantifiable mpox clade II DNA as measured during a prospective, wastewater monitoring program. These samples were chosen and tested for mpox clade Ib to test the specificity of the assay, given the reasonable assumption of the absence of clade Ib DNA in the samples because there had not been any clade I cases reported in the United States at that time. When originally measured during prospective monitoring, the concentration of clade II mpox in each sample (determined as described in detail by Boehm et al.[2]) ranged from non-detect to 19,315 cp/g wastewater solids (Figure S1). Samples were processed via the methods outlined below for pre-analytical processing and nucleic-acid extraction. Nucleic-acids were stored at -80°C for one to three years and underwent one freeze thaw to use as template in RT-PCR as described below.

**Wastewater sample collection for prospective study.** The process for wastewater collection has been provided elsewhere[2] previously and is summarized here. Samples were collected from 147 WWTPs across 40 states between 11/22/2025 and 1/31/2025 approximately three times per week. A full description of the WWTP locations, as well as the number of samples and the time during which they were collected is provided in Table S2. WWTP staff provided either "grab" samples from the primary clarifier or 24-hour composite samples from the headworks ("influent") (Table S2 provides sample type for each WWTP). Samples were then stored at 4°C and shipped to the laboratory on ice and processed to completion within 48 hours of receipt at the laboratory. We do not expect the time between sample collection and analysis to affect target quantification as our work and others[3–5] show limited decay of the short length nucleic-acid targets over weeks at 4°C. A total of 3,292 samples were collected and analyzed for this study.

**Pre-analytical processing and nucleic-acid extraction of wastewater samples.** The pre-analytical processing and nucleic-acid extraction of the samples has been described previously in detail[6] and is summarized here briefly. Solids are isolated from wastewater samples and re-suspended in a buffer at a low enough concentration so as to minimize inhibition, and homogenized. Samples were then subjected to nucleic-acid extraction and purification (Chemagic Viral DNA/RNA 300 Kit H96, PerkinElmer, Shelton, CT), and inhibitor removal (Zymo OneStep PCR Inhibitor Removal Kit, Irvine, CA). Dry weight was determined using another aliquot of solids and drying in an oven. Nucleic-acids from 6 to 10 aliquots of each sample were obtained and each used neat as template in 6 to 10 replicate droplet digital 1-step RT-PCR (dd-RT-PCR) reaction wells to measure mpox clade Ib DNA. The process includes negative and positive extraction and PCR controls, as well as a measure of recovery as determined from spiking exogenous bovine coronavirus (BCoV). Further details are described in a protocol on protocols.io[7,8] and in Boehm et al.[6].

**Droplet digital PCR.** Concentrations of the mpox clade Ib DNA marker in controls and samples were measured in multiplex droplet digital 1-step RT-PCR (dd-RT-PCR) reactions using an AutoDG Automated Droplet Generator (Bio-Rad, Hercules, CA), Mastercycler Pro (Eppendforf, Enfield, CT) thermocycler, and a QX600 Droplet Reader (Bio-Rad). RT-PCR was used for detection and quantification despite the target being genomic DNA because testing is implemented in multiplex with RNA viruses. The mpox clade Ib assay was run using a probe-mixing multiplexing approach using probes labeled with carboxy-X-rhodamine (ROX) and ATTO590 in multiplex with probe-based assays targeting hepatitis A, influenza B, SARS-CoV-2, RSV, influenza A and human norovirus GII as described in Boehm et al.[6] For samples processed on and after 27 January 2025, the probe was labeled with ROX alone.

Digital droplet RT-PCR was performed on 20 µl samples from a 22 µl reaction volume, prepared using 5.5 µl template, mixed with 5.5 µl of One-Step RT-ddPCR Advanced kit for Probes (catalog no. 1863021; Bio-Rad), 2.2 µl reverse transcriptase (RT), 1.1 µl dithiothreitol (DTT), and primers and probes at a final concentration of 900 nM and 250 nM, respectively. Droplets were generated using the AutoDG Automated Droplet Generator (Bio-Rad).

For the in vitro assay testing, a subset of samples was run without the RT step in singleplex (see above). The same mastermix was used, except that the RT and DTT were omitted from the mastermix and replaced with nuclease free water.

PCR was performed using Mastercycler Pro with the following protocol: reverse transcription at 50 °C for 60 min, enzyme activation at 95 °C for 5 min, 40 cycles with 1 cycle consisting of denaturation at 95 °C for 30 s and annealing and extension at 59 °C for 30 s, enzyme deactivation at 98 °C for 10 min, and then an indefinite hold at 4 °C. The ramp rate for temperature changes was set at 2 °C/s, and the final hold at 4 °C was performed for a minimum of 30 min to allow the droplets to stabilize.

Droplets were analyzed using the QX600 Droplet Reader (Bio-Rad). A well had to have over 10,000 droplets for inclusion in the analysis. Extraction and PCR positive and negative controls were run on each 96-well plate. Results from replicate wells were merged for analysis. Concentrations of the targets in wastewater samples are presented as copies per gram dry weight. For a sample to be scored as a positive, there had to be at least 3 positive droplets. The lowest measurable concentration is approximately 1000 copies/g dry weight (corresponds to three positive droplets). Errors are reported as standard deviations of the measurements as obtained from QX Manager Software (Bio-Rad, version 2.0).

**QA/QC.** All the negative controls and positive controls were negative and positive for all analyses in this study. Results for recovery of bovine coronavirus were reported elsewhere and indicate near uniform recovery[6].

For the prospective study, a total of 274 negative extraction controls and 274 no template controls (NTCs) were run evenly across 274 plates. The negative extraction controls and NTCs were run in 3 wells and the droplet counts were merged across wells (median total accepted droplet counts across negative extraction controls and NTCs were 39816 and 40202, respectively. There were 0 positive droplets in all 274 NTCs. There were 0 positive droplets in 273 of 274 negative extraction controls, and 1 positive droplet in 1 of the negative extraction controls.

In order for a sample to be scored as positive, it had to have 3 or more positive droplets across the merged replicate wells. Across the 3291 negative wastewater solids samples analyzed prospectively in this project, 3186 samples had 0 positive droplets, 84 samples had 1 positive droplet, and 20 samples had 2 positive droplets. The median total accepted droplet count across these samples was 115,050.

**Additional details related to the EMMI guidelines.** Across all the prospective samples analyzed in this study (n=3292), the average (standard deviation) number of partitions (droplets) for the across the 6 to 10 replicate wells was 119891 (40375). The volume of the partitions, as reported by the machine vendor is 0.00085 µL. The mean (standard deviation) of copies per partition for each target was $2.73 \times 10^{-7}$ ($1.69 \times 10^{-6}$) for mpox clade 1b gene. An example fluorescent plot from the QX600 (6 color reader), as well as the EMMI checklist is included in the Stanford Digital Repository with the deposited data (https://doi.org/10.25740/vk458br0887).

**Comparison between SARS-CoV-2 RNA measured in unstored and stored retrospective samples.** The samples used for the retrospective study were collected as part of an ongoing wastewater surveillance program. SARS-CoV-2 N gene concentrations were measured in them in a prospective manner, and the samples were not stored prior to the quantification of the N gene. Those measurements were previously published[6]. We compared those N gene measurements to the ones we obtained using the samples after they had been stored between 1 and 3 years to gain insight into potential nucleic-acid degradation. We found a significantly lower concentration between the N gene concentrations measured before and after storage in the 7 samples from 2022 (Figure S1 samples 1-7, Wilcoxon signed rank test $p = 0.02$), but no significant differences for the three samples from 2024 (Fig S1 samples 8-10, $p = 0.25$). For 2022 samples, the median ratio of the concentrations obtained after storage to those obtained before storage was 0.58 (interquartile range = 0.57 and 0.73), and for 2024 samples the median ratio was 1.57 (IQR = 1.54 and 1.99). Although this analysis was done using an RNA target and there was a statistically significant decrease in concentrations in the older set of samples, it suggests limited degradation in overall concentration and we suspect the same applies to the DNA.

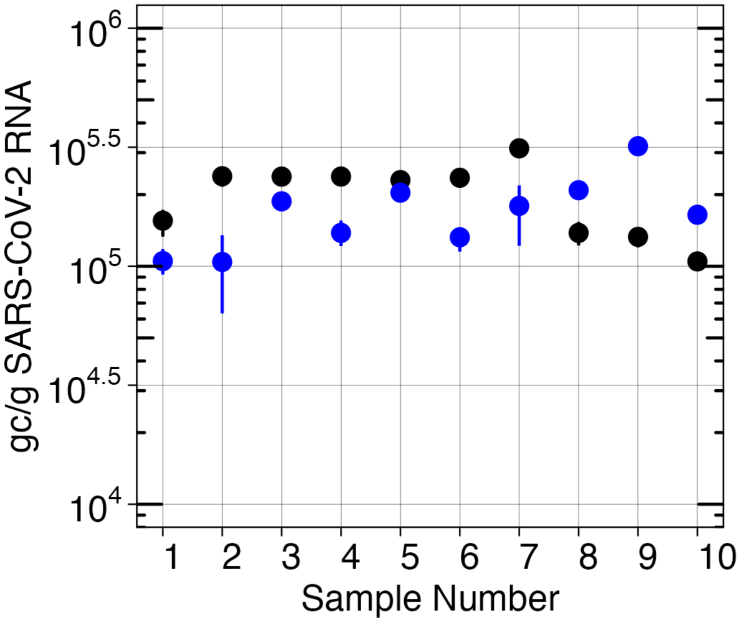


Figure S1. SARS-CoV-2 concentrations in units of gene copies per gram dry weight solids. Results in black are the concentrations generated when samples were originally tested at the time of collection, as reported by Boehm et al.[2]. Results generated at the same time as mpox clade Ib testing described in this paper are shown in blue. Error bars that are not visible are smaller than the plotted points. Samples numbers correspond to the following dates, in chronological order: 8/4 - 8/10/2022, 9/3/2024, 9/4/2024, 10/11/2024. Errors represent standard deviations.

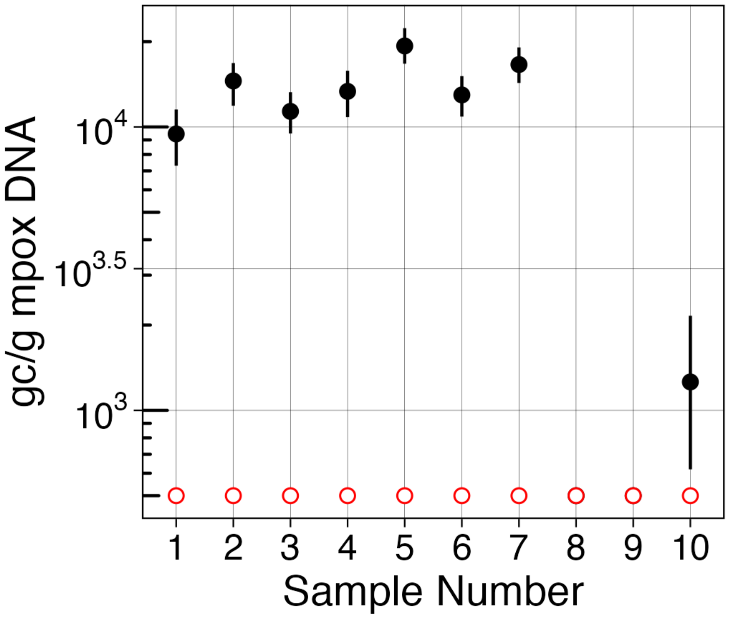


Figure S2. Mpox clade II concentrations in units of gene copies per gram dry weight solids and mpox clade Ib non-detects. Results for clade II are shown in black and reported by Boehm et al.[2]. Results for clade Ib are shown in red. Open circles indicate that the target was not detected. Samples numbers correspond to the following dates, in chronological order: 8/4 - 8/10/2022, 9/3/2024, 9/4/2024, 10/11/2024. If a black symbol is not visible, it is also non-detect and under the red symbol. Errors represent standard deviations.

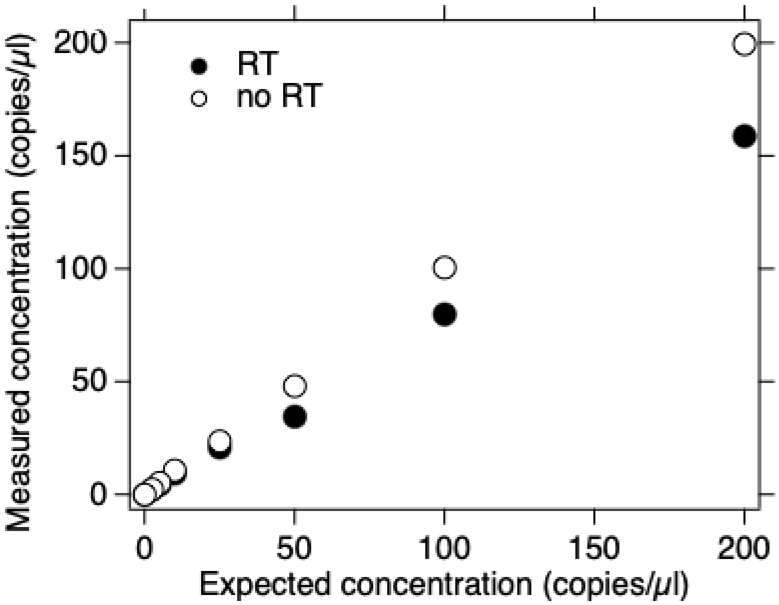

Fig S3. Concentrations of mpox clade Ib DNA in units of copies per μl of the reaction (total volume is 20 μl). Positive control DNA was spiked into nucleic-acids from wastewater known to be negative for the target and then quantified with and without an RT step (RT and no RT, respectively). Expected concentration is that expected in the sample given what was spiked into it. Measured is the actual measured concentration. Error bars are the standard deviations, but cannot be seen because they are smaller than the symbol.

| **Zeptometrix** | | **Twist** | **ATCC** |
|---|---|---|---|
| Parainfluenza 1<br>Parainfluenza 2<br>Parainfluenza 3<br>Parainfluenza 4<br>Influenza A H1N1pdm<br>Influenza AH1<br>Influenza AH3<br>Influenza B<br>Adenovirus 1<br>Adenovirus 3<br>Adenovirus 31<br>Rhinovirus Type 1A | RSV A<br>SARS-CoV-2<br>M. pneumoniae<br>C. pneumoniae<br>Metapneumovirus 8<br>Coronavirus HKU-1<br>Coronavirus 229E<br>Coronavirus NL63<br>Coronavirus OC43<br>B. parapertussis<br>B. pertussis<br>RSV B | Influenza B<br>Influenza A H1N1<br>Influenza A H3N2<br>Mpox Clade Ia<br>Mpox Clade II | Genomic SARS-CoV-2 gRNA (extracted)<br>Measles (entire RNA genome, synthetic)<br>Parvovirus B19 (fragments from VP1, VP2, and NS1 regions, synthetic) |

Table S1. Pathogens and genetic material used for in vitro specificity testing. Controls from Zeptometrix (Buffalo, NY) are inactivated pathogens. Controls from Twist (South San Francisco, CA) are whole genome synthetic nucleic acid. Controls from ATCC (Manassas, VA) include extracted and synthetic nucleic acids as described in the table.

Table S2. Each WWTP included in this study in each row. Solids were obtained the liquid samples by centrifugation as described elsewhere[6]. The number replicates indicates whether there are 6 or 10 replicates run as described elsewhere[6].

| State | Plant | Site Name | County | Population | First Sample Date | Last Sample Date | Sample Count | Sample Type | Number of replicates |
|---|---|---|---|---|---|---|---|---|---|
| AK | Anchorage, AK | John M. Asplund Water Pollution Control Facility | Anchorage | 220,000 | 12/17/24 | 1/30/25 | 19 | Liquids | 6 |
| AL | Bessemer, AL | Valley Creek Water Reclamation Facility | Jefferson | 225,000 | 12/13/24 | 1/31/25 | 21 | Liquids | 6 |
| AL | Cahaba River, Birmingham, AL | Cahaba River Water Reclamation Facility | Jefferson | 95,000 | 12/13/24 | 1/31/25 | 23 | Liquids | 6 |
| AL | Fultondale, AL | Five Mile Creek Water Reclamation Facility | Jefferson | 77,000 | 12/13/24 | 1/31/25 | 21 | Liquids | 6 |
| AL | Pinson, AL | Turkey Creek Water Reclamation Facility | Jefferson | 30,000 | 12/13/24 | 1/31/25 | 22 | Liquids | 6 |
| AL | Village Creek, Birmingham, AL | Village Creek Water Reclamation Facility | Jefferson | 200,000 | 12/13/24 | 1/31/25 | 21 | Liquids | 6 |
| AR | Harrison, AR | City of Harrison Wastewater Treatment Plant | Boone | 15,000 | 12/13/24 | 1/31/25 | 22 | Liquids | 6 |
| CA | Davis, CA | City of Davis Wastewater Treatment Plant | Yolo | 68,000 | 12/13/24 | 1/29/25 | 21 | Solids | 6 |
| CA | Fremont, CA | [Fremont Basin] - Raymond A. Boege Alvarado WWTP | Alameda | 229,476 | 12/12/24 | 1/30/25 | 13 | Liquids | 6 |
| CA | Gilroy, CA | South County Regional Wastewater Authority | Santa Clara | 110,338 | 11/23/24 | 1/31/25 | 70 | Solids | 10 |
| CA | Indio, CA | Valley Sanitary District | Riverside | 91,765 | 12/13/24 | 1/31/25 | 22 | Solids | 6 |

| State | Plant | Site Name | County | Population | First Sample Date | Last Sample Date | Sample Count | Sample Type | Number of replicates |
|---|---|---|---|---|---|---|---|---|---|
| CA | Lompoc, CA | Lompoc Regional Wastewater Reclamation Plant | Santa Barbara | 69,290 | 12/13/24 | 1/31/25 | 22 | Liquids | 6 |
| CA | Los Angeles County, CA | Joint Water Pollution Control Plant | Los Angeles | 3,500,000 | 12/15/24 | 1/29/25 | 21 | Liquids | 6 |
| CA | Los Angeles, CA | Hyperion Water Reclamation Plant (HWRP) | Los Angeles | 4,000,000 | 12/15/24 | 1/29/25 | 21 | Liquids | 6 |
| CA | Marina, CA | Monterey One Water - Regional Treatment Plant | Monterey | 262,000 | 12/13/24 | 1/31/25 | 20 | Liquids | 6 |
| CA | Merced, CA | Merced Wastewater Treatment Plant | Merced | 91,000 | 12/13/24 | 1/31/25 | 22 | Solids | 6 |
| CA | Napa, CA | Soscol Water Recycling Facility | Napa | 83,300 | 12/13/24 | 1/31/25 | 22 | Solids | 6 |
| CA | Novato, CA | Novato Sanitary District | Marin | 53,000 | 12/13/24 | 1/31/25 | 20 | Liquids | 6 |
| CA | Oceanside San Francisco, CA | Oceanside Water Pollution Control Plant | San Francisco | 250,000 | 11/23/24 | 1/31/25 | 69 | Solids | 10 |
| CA | Ontario, CA | Regional Water Recycling Plant No.1 (RP-1) | San Bernardino | 890,000 | 12/16/24 | 1/30/25 | 19 | Solids | 6 |
| CA | Palo Alto, CA | Palo Alto Regional Water Quality Control Plant | Santa Clara | 236,000 | 11/22/24 | 1/31/25 | 71 | Solids | 10 |
| CA | Paso Robles, CA | City of Paso Robles Wastewater Treatment Plant | San Luis Obispo | 31,037 | 12/16/24 | 1/30/25 | 20 | Solids | 6 |
| CA | Redwood City, CA | Silicon Valley Clean Water | San Mateo | 199,000 | 11/25/24 | 1/31/25 | 50 | Solids | 10 |
| CA | Riverside, CA | Riverside Water Quality Control Plant | Riverside | 350,000 | 12/13/24 | 1/31/25 | 22 | Liquids | 6 |

| State | Plant | Site Name | County | Population | First Sample Date | Last Sample Date | Sample Count | Sample Type | Number of replicates |
|---|---|---|---|---|---|---|---|---|---|
| CA | Sacramento, CA | Sacramento Regional Wastewater Treatment Plant | Sacramento | 1,480,000 | 11/23/24 | 1/31/25 | 70 | Solids | 10 |
| CA | San Diego, CA | E.W. Blom Point Loma Wastewater Treatment Plant | San Diego | 2,200,000 | 12/15/24 | 1/29/25 | 16 | Liquids | 6 |
| CA | San Jose, CA | San Jose-Santa Clara Regional Wastewater Facility | Santa Clara | 1,500,000 | 11/22/24 | 1/31/25 | 71 | Solids | 10 |
| CA | San Rafael, CA | Central Marin Sanitation Agency | Marin | 104,250 | 12/15/24 | 1/28/25 | 11 | Liquids | 6 |
| CA | Santa Cruz, CA | City of Santa Cruz WTF - City Influent | Santa Cruz | 160,000 | 12/12/24 | 1/30/25 | 22 | Solids | 6 |
| CA | Santa Rosa, CA | City of Santa Rosa, Laguna Treatment Plant | Sonoma | 230,000 | 12/15/24 | 1/29/25 | 21 | Solids | 6 |
| CA | Southeast San Francisco, CA | Southeast San Francisco | San Francisco | 750,000 | 11/23/24 | 1/31/25 | 66 | Solids | 10 |
| CA | Sunnyvale, CA | City of Sunnyvale Water Pollution Control Plant | Santa Clara | 153,000 | 11/23/24 | 1/31/25 | 69 | Solids | 10 |
| CA | Turlock, CA | Turlock Regional Water Quality Control Facility | Stanislaus | 86,000 | 12/13/24 | 1/31/25 | 22 | Liquids | 6 |
| CA | Vallejo, CA | Vallejo Flood and Wastewater District Wastewater Treatment Plant | Solano | 121,000 | 12/17/24 | 1/31/25 | 21 | Liquids | 6 |
| CA | Woodland, CA | Woodland Water Pollution Control Facility | Yolo | 59,000 | 12/11/24 | 1/31/25 | 17 | Liquids | 6 |

| State | Plant | Site Name | County | Population | First Sample Date | Last Sample Date | Sample Count | Sample Type | Number of replicates |
|---|---|---|---|---|---|---|---|---|---|
| CO | North, Parker, CO | Parker Water and Sanitation District North Water Reclamation Facility | Douglas | 35,000 | 12/17/24 | 1/30/25 | 16 | Liquids | 6 |
| CO | South, Parker, CO | Parker Water and Sanitation District South Water Reclamation Facility | Douglas | 25,000 | 12/17/24 | 1/30/25 | 16 | Liquids | 6 |
| CT | Stamford, CT | City of Stamford, Water Pollution Control Authority | Fairfield | 140,000 | 12/13/24 | 1/31/25 | 21 | Solids | 6 |
| DE | Seaford, DE | Seaford Wastewater Treatment Facility | Sussex | 13,172 | 12/13/24 | 1/31/25 | 22 | Solids | 6 |
| FL | Eastern, Orange County, FL | Eastern Water Reclamation Facility | Orange | 195,299 | 12/12/24 | 1/30/25 | 16 | Solids | 6 |
| FL | Jupiter, FL | Loxahatchee River Environmental Control District | Palm Beach | 90,000 | 12/16/24 | 1/31/25 | 21 | Liquids | 6 |
| FL | Key Biscayne, FL | MDWASD Central District WWTP | Miami-Dade | 829,725 | 12/15/24 | 1/28/25 | 21 | Liquids | 6 |
| FL | North Miami, FL | MDWASD North District WWTF | Miami-Dade | 776,150 | 12/13/24 | 1/31/25 | 22 | Liquids | 6 |
| FL | Northeast, Saint Petersburg, FL | Northeast Water Reclamation Facility | Pinellas | 89,847 | 12/13/24 | 1/31/25 | 19 | Liquids | 6 |
| FL | Northwest, Orange County, FL | Northwest Water Reclamation Facility | Orange | 66,690 | 12/12/24 | 1/30/25 | 16 | Solids | 6 |
| FL | Northwest, Saint Petersburg, FL | Northwest Water Reclamation Facility | Pinellas | 94,218 | 12/13/24 | 1/31/25 | 19 | Liquids | 6 |
| FL | South, Orange County, FL | South Water Reclamation Facility | Orange | 183,009 | 12/12/24 | 1/30/25 | 16 | Solids | 6 |

| State | Plant | Site Name | County | Population | First Sample Date | Last Sample Date | Sample Count | Sample Type | Number of replicates |
|---|---|---|---|---|---|---|---|---|---|
| FL | Southwest, Orange County, FL | Hamlin Water Reclamation Facility | Orange | 50,000 | 12/12/24 | 1/30/25 | 16 | Solids | 6 |
| FL | Southwest, Saint Petersburg, FL | Southwest Water Reclamation Facility | Pinellas | 47,790 | 12/13/24 | 1/31/25 | 20 | Liquids | 6 |
| FL | Tallahassee, FL | TPSmith Water Reclamation Facility | Leon | 212,065 | 12/12/24 | 1/30/25 | 22 | Liquids | 6 |
| GA | Big Creek, Roswell, GA | Big Creek Water Reclamation Facility | Fulton | 189,593 | 12/16/24 | 1/29/25 | 21 | Liquids | 6 |
| GA | College Park, GA | Camp Creek Water Reclamation Facility | Fulton | 73,821 | 12/11/24 | 1/29/25 | 22 | Liquids | 6 |
| GA | Columbus, GA | South Columbus Water Resources Facility | Muscogee | 278,000 | 12/16/24 | 1/29/25 | 14 | Solids | 6 |
| GA | Johns Creek, Roswell, GA | Johns Creek Environmental Campus | Fulton | 84,486 | 12/16/24 | 1/29/25 | 21 | Liquids | 6 |
| GA | Little River, Roswell, GA | Little River Water Reclamation Facility | Fulton | 12,818 | 12/16/24 | 1/29/25 | 21 | Liquids | 6 |
| GA | RM Clayton, Atlanta, GA | RM Clayton Water Reclamation Center | Fulton | 294,660 | 12/10/24 | 1/30/25 | 20 | Liquids | 6 |
| GA | South River, Atlanta, GA | South River Water Reclamation Center | Fulton | 105,160 | 12/10/24 | 1/30/25 | 20 | Liquids | 6 |
| GA | Utoy Creek, Atlanta, GA | Utoy Creek Water Reclamation Center | Fulton | 70,887 | 12/10/24 | 1/30/25 | 20 | Liquids | 6 |
| HI | Honouliuli, Honolulu, HI | Honouliuli Wastewater Treatment Plant | Honolulu | 300,000 | 12/16/24 | 1/29/25 | 21 | Liquids | 6 |

| State | Plant | Site Name | County | Population | First Sample Date | Last Sample Date | Sample Count | Sample Type | Number of replicates |
|---|---|---|---|---|---|---|---|---|---|
| HI | Sand Island, Honolulu, HI | Sand Island Wastewater Treatment Plant | Honolulu | 390,000 | 12/16/24 | 1/29/25 | 21 | Liquids | 6 |
| IA | Clinton, IA | City of Clinton | Clinton | 29,300 | 12/13/24 | 1/31/25 | 22 | Solids | 6 |
| IA | Coralville, IA | Coralville Wastewater Treatment Facility | Johnson | 23,000 | 12/13/24 | 1/31/25 | 20 | Liquids | 6 |
| IA | Marshalltown, IA | City of Marshalltown Water Pollution Control Plant | Marshall | 27,400 | 12/12/24 | 1/30/25 | 22 | Liquids | 6 |
| IA | Muscatine, IA | Muscatine STP | Muscatine | 24,400 | 12/12/24 | 1/30/25 | 23 | Solids | 6 |
| IA | Ottumwa, IA | Ottumwa WPCF | Wapello | 25,529 | 12/13/24 | 1/31/25 | 22 | Liquids | 6 |
| ID | Coeur d'Alene, ID | City of Coeur d'Alene Water Resource Recovery Facility | Kootenai | 50,540 | 12/13/24 | 1/31/25 | 19 | Solids | 6 |
| ID | Lander Street, Boise, ID | Lander Street Water Renewal Facility | Ada | 108,556 | 12/13/24 | 1/31/25 | 22 | Liquids | 6 |
| ID | West Boise, ID | West Boise Water Renewal Facility | Ada | 186,901 | 12/13/24 | 1/31/25 | 21 | Liquids | 6 |
| IL | Glen Ellyn, IL | Glenbard Wastewater Authority | DuPage | 86,000 | 12/16/24 | 1/30/25 | 19 | Solids | 6 |
| IL | Wheaton, IL | Wheaton Sanitary District | DuPage | 63,000 | 12/13/24 | 1/31/25 | 22 | Solids | 6 |
| IN | Bloomington, IN | Dillman Road WWTP | Monroe | 56,090 | 12/16/24 | 1/30/25 | 16 | Liquids | 6 |
| IN | Carmel, IN | City of Carmel WWTP | Hamilton | 86,000 | 12/13/24 | 1/31/25 | 23 | Solids | 6 |
| IN | Downtown, Jeffersonville, IN | Jeffersonville Downtown WWTP | Clark | 25,000 | 12/13/24 | 1/31/25 | 22 | Liquids | 6 |
| IN | North, Jeffersonville, IN | North Water Reclamation Facility | Clark | 25,000 | 12/13/24 | 1/31/25 | 22 | Liquids | 6 |

| State | Plant | Site Name | County | Population | First Sample Date | Last Sample Date | Sample Count | Sample Type | Number of replicates |
|---|---|---|---|---|---|---|---|---|---|
| IN | South Bend, IN | City of South Bend Wastewater Treatment Plant | St. Joseph | 130,000 | 12/12/24 | 1/30/25 | 22 | Liquids | 6 |
| KS | Kaw Point, Kansas City, KS | Municipal Wastewater Treatment Plant No. 1 (Kaw Point) | Wyandotte | 90,000 | 12/10/24 | 1/30/25 | 19 | Solids | 6 |
| KS | Lawrence, KS | Lawrence Kansas River Wastewater Treatment Facility | Douglas | 80,000 | 12/13/24 | 1/31/25 | 19 | Solids | 6 |
| KS | P20, Kansas City, KS | Kansas City Treatment Plant #20 | Wyandotte | 35,000 | 12/10/24 | 1/30/25 | 19 | Solids | 6 |
| KS | Salina, KS | Salina Wastewater Treatment Plant | Saline | 47,000 | 12/13/24 | 1/31/25 | 22 | Solids | 6 |
| KS | Wolcott, Kansas City, KS | Wolcott Wastewater Treatment Facility | Wyandotte | 15,000 | 12/10/24 | 1/30/25 | 12 | Liquids | 6 |
| KY | Louisville, KY | Morris Forman Water Quality Treatment Center | Jefferson | 423,913 | 12/17/24 | 1/29/25 | 12 | Solids | 6 |
| LA | East Bank, New Orleans, LA | SWBNO East Bank Wastewater Treatment Plant | Orleans | 333,406 | 12/13/24 | 1/29/25 | 18 | Liquids | 6 |
| LA | West Bank, New Orleans, LA | SWBNO West Bank Wastewater Treatment Plant | Orleans | 50,591 | 12/13/24 | 1/31/25 | 21 | Liquids | 6 |
| MA | Boston, MA | Deer Island Treatment Plant | Suffolk | 2,400,000 | 12/13/24 | 1/31/25 | 21 | Solids | 6 |
| MA | Millbury, MA | Upper Blackstone Clean Water | Worcester | 250,000 | 12/12/24 | 1/30/25 | 19 | Liquids | 6 |
| MD | Hagerstown, MD | Hagerstown Wastewater Treatment Plant | Washington | 90,000 | 12/13/24 | 1/31/25 | 22 | Liquids | 6 |

| State | Plant | Site Name | County | Population | First Sample Date | Last Sample Date | Sample Count | Sample Type | Number of replicates |
|---|---|---|---|---|---|---|---|---|---|
| MD | Hollywood, MD | Marlay Taylor Water Reclamation Facility | St. Mary's | 55,000 | 12/13/24 | 1/29/25 | 13 | Liquids | 6 |
| ME | Bangor, ME | City of Bangor Wastewater Treatment Plant | Penobscot | 40,000 | 12/16/24 | 1/30/25 | 18 | Solids | 6 |
| ME | Lewiston, ME | Lewiston Aburn Water Pollution Control Authority | Androscoggin | 60,000 | 12/13/24 | 1/30/25 | 20 | Liquids | 6 |
| ME | Portland, ME | Portland Water District (East End Wastewater Treatment Facility) | Cumberland | 65,000 | 12/13/24 | 1/31/25 | 20 | Liquids | 6 |
| MI | Ann Arbor, MI | City of Ann Arbor Wastewater Treatment Plant | Washtenaw | 125,000 | 12/13/24 | 1/31/25 | 22 | Liquids | 6 |
| MI | Jackson, MI | Jackson Wastewater Treatment Plant | Jackson | 90,000 | 12/12/24 | 1/30/25 | 22 | Solids | 6 |
| MI | Jenison, MI | Grandville Clean Water Plant | Ottowa | 75,000 | 12/13/24 | 1/31/25 | 22 | Solids | 6 |
| MI | Mt. Pleasant, MI | Mt. Pleasant WRRF | Isabella | 21,690 | 12/13/24 | 1/29/25 | 20 | Liquids | 6 |
| MI | Traverse City, MI | Traverse City Regional Waste Water Treatment Plant | Grand Traverse | 30,623 | 12/16/24 | 1/30/25 | 20 | Liquids | 6 |
| MI | Warren, MI | City of Warren Wastewater Treatment Plant | Macomb | 140,000 | 12/16/24 | 1/30/25 | 19 | Liquids | 6 |
| MN | Mankato, MN | City of Mankato Water Resource Recovery Facility (WRRF) | Blue Earth | 70,000 | 12/16/24 | 1/31/25 | 21 | Solids | 6 |
| MN | Red Wing, MN | Red Wing Wastewater Treatment Facility | Goodhue | 16,000 | 12/13/24 | 1/31/25 | 22 | Solids | 6 |

| State | Plant | Site Name | County | Population | First Sample Date | Last Sample Date | Sample Count | Sample Type | Number of replicates |
|---|---|---|---|---|---|---|---|---|---|
| MN | Rochester, MN | City Of Rochester MN Water Reclamation Plant | Olmstead | 120,000 | 12/13/24 | 1/31/25 | 22 | Solids | 6 |
| MN | St. Cloud, MN | St. Cloud Nutrient, Energy and Water Recovery Facility | Stearns | 120,000 | 12/13/24 | 1/31/25 | 22 | Liquids | 6 |
| MS | Gautier, MS | 2C-Gautier POTW | Jackson | 19,008 | 12/15/24 | 1/28/25 | 21 | Liquids | 6 |
| MS | Pascagoula Moss Point, MS | 7 C- Pascagoula Moss Point POTW | Jackson | 34,333 | 12/8/24 | 1/30/25 | 24 | Liquids | 6 |
| NC | Kinston, NC | Johnnie Mosley Regional Water Reclamation Facility | Lenoir | 25,000 | 12/13/24 | 1/31/25 | 19 | Liquids | 6 |
| NC | Wilson, NC | City of Wilson - Hominy Creek Water Reclamation Facility | Wilson | 50,000 | 12/13/24 | 1/31/25 | 19 | Solids | 6 |
| NC | Winston-Salem, NC | Archie Elledge WWTP | Forsyth | 92,000 | 12/13/24 | 1/31/25 | 16 | Liquids | 6 |
| NE | Northeast, Lincoln, NE | Northeast Water Resource Recovery Facility | Lancaster | 60,000 | 12/13/24 | 1/31/25 | 22 | Liquids | 6 |
| NE | Theresa Street, Lincoln, NE | Theresa Street Water Resource Recovery Facility | Lancaster | 240,000 | 12/13/24 | 1/31/25 | 22 | Liquids | 6 |
| NH | Dover, NH | City of Dover Wastewater Treatment Facility | Strafford | 30,000 | 12/16/24 | 1/30/25 | 20 | Liquids | 6 |
| NJ | Belmar, NJ | South Monmouth Regional Sewerage Authority | Monmouth | 52,672 | 12/13/24 | 1/31/25 | 22 | Solids | 6 |
| NJ | Bridgeton, NJ | Cumberland County Utilities Authority | Cumberland | 50,000 | 12/13/24 | 1/31/25 | 19 | Liquids | 6 |

| State | Plant | Site Name | County | Population | First Sample Date | Last Sample Date | Sample Count | Sample Type | Number of replicates |
|---|---|---|---|---|---|---|---|---|---|
| NJ | Bridgewater, NJ | The Somerset Raritan Valley Sewerage Authority | Somerset | 130,000 | 12/13/24 | 1/31/25 | 14 | Liquids | 6 |
| NJ | Newark, NJ | Passaic Valley Sewerage Commission | Essex | 1,500,000 | 12/23/24 | 1/29/25 | 13 | Solids | 6 |
| NJ | Oakhurst, NJ | Township of Ocean Sewerage Authority | Monmouth | 50,000 | 12/13/24 | 1/31/25 | 21 | Liquids | 6 |
| NJ | Union Beach, NJ | Bayshore Regional Sewerage Authority | Monmouth | 100,000 | 12/13/24 | 1/31/25 | 22 | Solids | 6 |
| NV | Las Vegas, NV | Clark County Water Reclamation District (CCWRD) Flamingo Water Resource Center (FWRC) | Clark | 990,000 | 12/13/24 | 1/31/25 | 22 | Liquids | 6 |
| NY | Ithaca, NY | Ithaca Area Wastewater Treatment Facility | Tompkins | 90,000 | 12/16/24 | 1/29/25 | 12 | Liquids | 6 |
| NY | Oswego, NY | City of Oswego Wastewater Treatment Plant | Oswego | 30,000 | 12/13/24 | 1/31/25 | 21 | Liquids | 6 |
| OH | Akron, OH | Akron Water Reclamation Facility | Summit | 365,000 | 12/13/24 | 1/31/25 | 21 | Solids | 6 |
| OH | Youngstown, OH | City of Youngstown Wastewater Treatment Plant | Mahoning | 174,000 | 12/13/24 | 1/30/25 | 22 | Solids | 6 |
| PA | Chester, PA | DELCORA Western Regional Treatment Plant | Delaware | 220,000 | 12/12/24 | 1/30/25 | 19 | Liquids | 6 |
| PA | Harrisburg, PA | Capital Region Water AWTF | Dauphin | 125,000 | 12/13/24 | 1/31/25 | 22 | Liquids | 6 |

| State | Plant | Site Name | County | Population | First Sample Date | Last Sample Date | Sample Count | Sample Type | Number of replicates |
|---|---|---|---|---|---|---|---|---|---|
| SD | Yankton, SD | City of Yankton Wastewater Treatment Facility | Yankton | 20,000 | 12/12/24 | 1/30/25 | 22 | Liquids | 6 |
| TN | Chattanooga, TN | Moccasin Bend WWTP | Hamilton | 400,000 | 12/11/24 | 1/31/25 | 19 | Liquids | 6 |
| TN | Memphis, TN | M.C. Stiles Wastewater Treatment Facility | Shelby | 300,000 | 12/12/24 | 1/30/25 | 22 | Liquids | 6 |
| TX | Dallas Central, Dallas, TX | DCWT Dallas | Dallas | 270,000 | 12/13/24 | 1/30/25 | 20 | Liquids | 6 |
| TX | Gainesville, TX | City of Gainesville Wastewater Treatment Plant | Cooke | 17,300 | 12/13/24 | 1/31/25 | 22 | Liquids | 6 |
| TX | Garland, TX | City of Garland Rowlett Creek WWTP | Dallas | 200,000 | 12/13/24 | 1/31/25 | 19 | Solids | 6 |
| TX | Hollywood Road, Amarillo, TX | Hollywood Road WWTP | Potter | 60,000 | 12/15/24 | 1/28/25 | 21 | Liquids | 6 |
| TX | River Road, Amarillo, TX | River Road WWTP | Potter | 140,000 | 12/15/24 | 1/28/25 | 21 | Liquids | 6 |
| TX | South, Laredo, TX | South Laredo WWTP | Webb | 120,000 | 12/13/24 | 1/31/25 | 19 | Liquids | 6 |
| TX | Southside, Dallas, TX | Southside Wastewater Treatment Plant (City of Dallas) | Dallas | 421,700 | 12/12/24 | 1/30/25 | 17 | Liquids | 6 |
| TX | Sunnyvale, TX | Duck Creek Wastewater Treatment Plant | Dallas | 186,000 | 12/12/24 | 1/30/25 | 22 | Solids | 6 |
| TX | White Rock Central, Dallas, TX | DCWT White Rock | Hunt | 630,000 | 12/13/24 | 1/30/25 | 21 | Liquids | 6 |
| TX | Wichita Falls, TX | Wichita Falls Resource Recovery Facility | Wichita | 90,000 | 12/13/24 | 1/31/25 | 22 | Solids | 6 |

| State | Plant | Site Name | County | Population | First Sample Date | Last Sample Date | Sample Count | Sample Type | Number of replicates |
|---|---|---|---|---|---|---|---|---|---|
| TX | Woodlands SJRA WWTF No. 1, TX | SJRA WWTF No.1 | San Jacinto | 65,000 | 12/13/24 | 1/31/25 | 22 | Liquids | 6 |
| TX | Woodlands SJRA WWTF No. 2, TX | SJRA WWTF No.2 | San Jacinto | 70,000 | 12/13/24 | 1/31/25 | 22 | Liquids | 6 |
| TX | Zacate Creek, Laredo, TX | Zacate Creek WWTP | Webb | 140,000 | 12/13/24 | 1/31/25 | 19 | Liquids | 6 |
| UT | Central Salt Lake Valley, UT | Central Valley Water Reclamation Facility | Salt Lake | 600,000 | 12/13/24 | 1/31/25 | 22 | Solids | 6 |
| UT | Provo, UT | Provo City Water Reclamation Facility | Utah | 115,000 | 12/17/24 | 1/29/25 | 14 | Solids | 6 |
| VA | Aquia, Stafford, VA | Aquia Wastewater Treatment Facility | Stafford | 100,000 | 12/16/24 | 1/31/25 | 13 | Solids | 6 |
| VA | Little Falls Run, Stafford, VA | Little Falls Run Wastewater Treatment Facility | Stafford | 50,000 | 12/16/24 | 1/31/25 | 13 | Solids | 6 |
| VT | Essex Junction, VT | City of Essex Junction Wastewater Treatment Facility | Chittenden | 30,000 | 12/13/24 | 1/31/25 | 22 | Solids | 6 |
| VT | Montpelier, VT | Montpelier Water Resource Recovery Facility | Washington | 10,100 | 12/16/24 | 1/30/25 | 19 | Solids | 6 |
| VT | South Burlington, VT | South Burlington-Airport Parkway WWTF | Chittenden | 16,000 | 12/17/24 | 1/30/25 | 11 | Liquids | 6 |
| WA | Snohomish, WA | City of Snohomish Wastewater Treatment Plant | Snohomish | 10,150 | 12/13/24 | 1/31/25 | 22 | Liquids | 6 |
| WI | Wausau, WI | Wausau Waterworks Wastewater Treatment Facility | Marathon | 44,000 | 12/11/24 | 1/31/25 | 23 | Solids | 6 |

| State | Plant | Site Name | County | Population | First Sample Date | Last Sample Date | Sample Count | Sample Type | Number of replicates |
|---|---|---|---|---|---|---|---|---|---|
| WV | Wheeling, WV | City of Wheeling, Water Pollution Control Division | Ohio | 100,000 | 12/13/24 | 1/31/25 | 22 | Liquids | 6 |

### References


(1) Schuele, L.; Masirika, L. M.; Udahemuka, J. C.; Siangoli, F. B.; Mbiribindi, J. B.; Ndishimye, P.; Aarestrup, F. M.; Koopmans, M.; Munnink, B. B. O.; Molenkamp, R.; Group, G. M. Real-Time PCR Assay to Detect the Novel Clade Ib Monkeypox Virus, September 2023 to May 2024. *Eurosurveillance* **2024**, *29* (32), 2400486. https://doi.org/10.2807/1560-7917.ES.2024.29.32.2400486.

(2) Boehm, A. B.; Wolfe, M. K.; Bidwell, A. L.; Zulli, A.; Chan-Herur, V.; White, B. J.; Shelden, B.; Duong, D. Human Pathogen Nucleic Acids in Wastewater Solids from 191 Wastewater Treatment Plants in the United States. *Scientific Data* **2024**, *11* (1), 1141. https://doi.org/10.1038/s41597-024-03969-8.

(3) Roldan-Hernandez, L.; Graham, K. E.; Duong, D.; Boehm, A. B. Persistence of Endogenous SARS-CoV-2 and Pepper Mild Mottle Virus RNA in Wastewater-Settled Solids. *ACS EST Water* **2022**. https://doi.org/10.1021/acsestwater.2c00003.

(4) Zhang, M.; Roldan-Hernandez, L.; Boehm, A. B. Persistence of Human Respiratory Viral RNA in Wastewater-Settled Solids. *Applied and Environmental Microbiology* **2024**, *90* (4), e02272-23. https://doi.org/10.1128/aem.02272-23.

(5) Muirhead, A.; Zhu, K.; Brown, J.; Basu, M.; Brinton, M. A.; Costa, F.; Hayat, M. J.; Stauber, C. E. Zika Virus RNA Persistence in Sewage. *Environ. Sci. Technol. Lett.* **2020**. https://doi.org/10.1021/acs.estlett.0c00535.

(6) Boehm, A. B.; Wolfe, M. K.; Bidwell, A. L.; Zulli, A.; Chan-Herur, V.; White, B. J.; Shelden, B.; Duong, D. Human Pathogen Nucleic Acids in Wastewater Solids from 191 Wastewater Treatment Plants in the United States. *Sci Data* **2024**, *11* (1), 1141. https://doi.org/10.1038/s41597-024-03969-8.

(7) Topol, A.; Wolfe, M.; White, B.; Wigginton, K.; Boehm, A. B. *High Throughput pre-analytical processing of wastewater settled solids for SARS-CoV-2 RNA analyses*. protocols.io. https://www.protocols.io/view/high-throughput-pre-analytical-processing-of-waste-b2kmqcu6 (accessed 2022-07-25).

(8) Topol, A.; Wolfe, M.; Wigginton, K.; White, B.; Boehm, A. *High Throughput RNA Extraction and PCR Inhibitor Removal of Settled Solids for Wastewater Surveillance of S...* protocols.io. https://www.protocols.io/view/high-throughput-rna-extraction-and-pcr-inhibitor-r-b2mkqc4w (accessed 2022-07-25).
